# Community participation and community engagement in the response to the Zika-Virus-Outbreak in Latin America 2015-2019 – A Scoping Review

**DOI:** 10.1101/2022.06.27.22276908

**Authors:** Sonja Nachtnebel, Ruth Kutalek

## Abstract

**Introduction:** Zika virus infection during pregnancy is often associated with the occurrence of numerous neurologic malformations in new-born babies. Usually, the transmission of the Zika virus happens through the bite of an infected mosquito of the Aedes genus but might as well occur through sexual contact or blood transfusions. Currently, there is no approved specific treatment, medication-based prophylaxis or vaccine available against Zika virus infection, thus prevention measures play an important role in combating the spread of this infectious disease. This scoping review aims to collect data about public health programmes in context of the Zika virus epidemic in Latin America that employ the principles of a community-based participatory research.

**Methods:** Three scientific databases (Medline, Cochrane Library and Scopus) were screened for relevant literature and additionally, official websites of health organizations such as WHO, UNICEF, ECDC, CDC and PAHO were included in the search. The review was conducted following the “Preferred Reporting Items for Scoping Reviews and Meta-Analysis extension for Scoping Reviews” (PRISMA-ScR) checklist. The authors developed a data-charting map to collect relevant information from each publication to analyse the content.

**Results:** Overall, 46 papers were included in the review. Each of the studies contained at least one of the following indicators: actual participation or involvement in a public health intervention, assessment of knowledge or awareness degree and preventative actions, assessment of disease perception or behaviour changes. Key topics were divided into four broad categories: 1) Mosquito vector control initiatives, 2) Contraception and reproductive healthcare, 3) Family support programmes and 4) Assessment of knowledge, perception and barriers.

**Conclusions:** Through engaging local communities, especially vulnerable groups, awareness of risks associated with a Zika virus infection can be raised and enable people to protect themselves. The current work demonstrates that community engagement is an essential strategy to implement preventive measures and improve health-outcomes.

## Introduction

The existence of the Zika virus was discovered in a rhesus macaque of the Ziika-Forest in Uganda in 1947 as result of investigations about the Yellow-fever virus and detected in humans a few years later in 1952. It belongs to the virus family Flaviviridae and is primarily transmitted to humans through the bite of infected mosquitoes of the daytime-active Aedes genus, particularly the subspecies Aedes aegypti and Aedes albopictus, which appears in tropical and subtropical regions of all continents. Additionally, the virus can be passed through sexual intercourse, blood transfusions, organ transplantation and perinatal. **[1, 2, 3, 4]** Despite its early identification, larger outbreaks of Zika virus occurred more than five decades later, in Yap Island 2007, and 2013 in French Polynesia. On the American continent the first confirmed Zika virus cases were reported in 2015. **[2, 5, 6]** After a sudden increase of the number of new-borns with congenital microcephaly and other neurological disorders was reported in the northern region of Brazil, the WHO subsequently declared the Zika virus outbreak a Public Health Emergency of International Concern (PHEIC) in February 2016. **[7]** The peak of the Zika virus epidemic reached its maximum in 2016. Until the end of the year, 651.690 suspected and more than 200.000 confirmed cases of Zika virus infections had been reported in the Americas, and about 3.000 cases of Congenital Zika Syndrome had been documented in Brazil only. **[5, 8]** By 2019 the Zika virus affected 86 countries and territories in North-Central- and South America, Asia and Africa **[5, 10]**. In Europe, two autochthonous cases of Zika virus appeared in early August 2019 in Southern France. The “Asian tiger mosquito” Aedes albopictus, which is less competent than the Aedes aegypti mosquito, became domestic in the meantime but environmental factors like weather conditions during the winter season were not suitable for further transmission. **[5, 10]** Currently, Zika is not considered a Public Health Emergency of International Concern anymore because of decreasing numbers in the last couple of years. **[5, 11, 12, 13]** Nevertheless, Zika is still endemic in Latin America and the Caribbean and is considered as an ongoing health threat, which hits especially vulnerable population such as pregnant women and women of reproductive age. Furthermore, global warming might lead to an increased spread of infectious diseases like Zika in the future. **[3, 5, 12, 14]** Most cases of Zika Virus Infection are asymptomatic or show a mild, self-limiting course of disease with unspecific symptoms like subfebrile temperatures, skin rash (maculopapular exanthema), arthralgia, conjunctivitis, headache, muscle pain and malaise, which might occur after an incubation period of three to fourteen days. The disease usually lasts two to seven days without the need for inpatient hospitalization. **[2, 3, 15]** After the increased incidence of new-born babies suffering from microcephaly and brain abnormalities at the end of 2015, further investigations found that a Zika virus infection in-utero – especially in the first trimester of pregnancy – might cause profound neurological consequences for the unborn child such as microcephaly, brain atrophy, ocular abnormalities, and hearing loss. **[1-4, 6, 15]** An acute Zika virus infection can be diagnosed with blood or urine tests one week after the infection through serological screening. Zika virus RNA can be found in body fluids like semen, saliva, cerebrospinal fluid, vaginal or cervical secretions and breast milk. The detection of viral RNA through usage of reverse-transcriptase-polymerase-chain-reaction (RT-PCR) assay delivers a rapid test result after two or three days of a Zika virus infection. **[3-6, 16, 17]** A specific antiviral treatment or vaccine is still not available and there is no known way to prevent a mother-to-child transmission of Zika virus during pregnancy yet. **[16]** Thus, the primary focus of public healthcare is set on exposure prophylaxis, mosquito control, sexual and reproductive health care and health education as preventive work in populations living in Zika-endemic regions. **[13, 14, 16]**

### Community engagement

As the World Health Organization recommends, risk communication and community engagement are at the centre of any public health intervention, especially in emergencies. **[18]** A “community” is defined as a minimum social group or network, which shares common values, the same habitats, lives or works under the same conditions, represents joint interests and pursues a common goal. Communities do not have to be geographically connected. **[13]** According to WHO, ECDC and UNICEF, community engagement builds on the three principles of human rights-based approach, community-based approach, quality and accountability. More detailed, community engagement is an approach that involves community residents and all official partners like government officials or health authorities into the assessment, planning, development, implementation, monitoring and evaluation processes and policies of a public health project. Community responsibility and awareness of subjects that need to be addressed, for instance the risks associated with infectious threats like Zika, should be raised. **[13, 14, 19-23]** Intervention strategies to manage the issues that affect the at-risk populations’
s lives should be developed and applied building on local capacities and adapted to the local context, considering diversity within the community like gender, age, ethnicity, religious beliefs, physical abilities and socioeconomic status. Most notably community engagement increases participation, ownership, communication and empowers social groups to take their own actions and should enhance existing capacities. An intersectoral cooperation at national, state and municipal level and a coordinated implementation of community-based initiatives could strengthen environmental management, improve residents’ knowledge and health status, reduce inequalities and enhance health care. **[13, 14, 39-43]**

Poverty, insufficient capability to respond to outbreaks and health inequity are the main risk factors for poor health and increase the burden of economic and social costs. **[22-28]** Insufficient access to healthcare services and information affects people’
ss health status, might lead to an increased risk of Zika virus transmission and could worsen the consequences of an epidemic outbreak. **[22, 25, 26]** As poverty affects reproductive health as well, disadvantaged families with low socioeconomic status, such as low educational background, income or unemployment, present the most vulnerable groups. Particularly pregnant women, adolescents and women of childbearing age and children are affected by a Zika virus infection. A child with CZS requires potentially lifelong assistance, intensive therapy and medical treatment and thus a higher need for quality healthcare services. The long-term consequences of health effects related to the Zika virus infection in children imply not only a heavy financial but also a psychological and social burden for affected families. **[23, 25, 29-31]** For this reason, mosquito vector control, access to reproductive healthcare services during pregnancy, appropriate fertility control, safe abortions, and sexual health education to prevent unintended pregnancies and sexually transmitted diseases in general are an important public health issue in the response to the Zika virus outbreak. **[23, 26, 32-38]** The application of community engagement principles targeting mosquito vector control and health education might improve maintenance of health outcomes, and therefore be a cost-effective approach in economic, political and social aspects. **[13, 14, 39-43]**

In the light of these challenges, a scoping review was performed to survey the available, scientific literature on community engagement and community participation strategies that have been established in context of the Zika-Virus epidemic in Latin America from 2015 to 2019. The review aims to assess community-based initiatives and programs, prevention activities of communities and identify barriers to healthcare access of vulnerable groups.

## Methods

A scoping review provides a broad overview of existing research evidence from different sources, might identify research gaps and give recommendations for future investigations. **[45, 46]** The review follows the five-stage methodological framework of Arksey and O’
sMalley, that includes (1) identifying the research question, (2) identifying relevant studies, (3) selecting the studies according to inclusion criteria, (4) charting and interpreting the data and (5) summarizing and reporting the results. **[45]** The authors applied “The Mixed Methods Appraisal Tool” (MMAT), which allows to assess studies of different designs (qualitative and quantitative studies, randomised controlled trials, quantitative non-randomised trials, descriptive quantitative studies, mixed methods studies) and therefore achieves critical appraisal of the studies. Furthermore, the scoping review was conducted based on the “Preferred Reporting Items for Scoping Reviews and Meta-Analysis extension for Scoping Reviews” (PRISMA-ScR) checklist. **[46]**

### Search strategy and study selection

An extensive literature search was performed to identify all relevant studies and available literature on the subject by searching three scientific electronic databases PubMed, Cochrane Library and Scopus. These databases were chosen because of comprehensive coverage on the subject due to a lot of evidence-based research results. Moreover, official health websites of the WHO, PAHO, Centres for Disease Control and Prevention, UNICEF, ECDC and sites of local health ministries were searched for grey literature.

The search string had three prior elements to gain detailed information, indicating whether the selected articles 1) reported initiatives, campaigns or programs in context of the Zika virus; 2) assessed knowledge, attitudes and individual preventive behaviour against Zika virus; and 3) identified barriers and health care access of vulnerable population in Latin America and the Caribbean countries. Hence, the search strategy consisted of the most relevant combination of key words as follows: “Zika virus” AND “community” or “perception” or “engagement” or “attitude” or “participation” or “prevention” or “education” or “mobilisation” or “vector control” or “Aedes” or “health access” or “empowerment”, adjusting search strings and MeSH terms.

### Inclusion criteria

Studies were eligible for inclusion when they were written in English or Spanish, published between January 2015 and May 2020 and contained a report, description or analysis of educational interventions, risk communication strategies, community engagement programmes, Aedes aegypti and vector control strategies, experiences of access to health care services, perspectives and knowledge about Zika virus infection, targeting pregnant women and women of reproductive age in particular. Documents were excluded when they merely reported incidence, seroprevalence, epidemiologic evidence or structural biology of Zika virus, studies published before the year 2015 and studies which were not performed in Latin America or the Caribbean.

### Data extracting and charting data

We developed a specific table to collect and extract data from each publication following the recommendations of the PRISMA guidelines. Information was categorized and compiled based on the country where the study had been conducted, year of publication and study design. **[46]** Additionally, we performed a content analysis of all included studies, summarized and extracted the data relating to the aim of the study in note form, the components of intervention or community engagement, main outcomes of the study and challenges or recommendations for future research.

## Results

A total of 9946 potentially relevant articles was obtained. After screening title, abstract and cross-references and removing duplicates, 246 studies could be identified, which possibly met the inclusion criteria. The full text of the eligible 246 published articles was reviewed and the study selection process finally resulted in 46 papers included in the review as seen from the flow diagram in Figure 1. Of the 46 included documents, 45 were published in English and one in Spanish. Of the peer-reviewed documents there was heterogeneity in study designs (based on MMAT classification), which included twenty qualitative studies **[8, 21-24, 26, 29, 32, 35, 36, 38, 47-54, 56]**, nine cross-sectional mixed method studies **[27, 30, 31, 37, 57-61]**, one cluster randomised controlled trial **[62]**, four descriptive studies **[63-66]** two reviews **[67, 68]**, three surveys **[34, 69, 70]**, one cohort study **[33]**, one pre-post design study **[71]**, and, four opinion and commentary essays **[28, 72-74]**. The majority of the studies was performed in Brazil (n=10) and in Puerto Rico (n=6), moreover in Mexico (n=5) Colombia (n= 4), Peru (n=4), Dominican Republic (n=3), Guatemala (n=2), Honduras (n=2), Ecuador (n=2), Cuba (n=2), Nicaragua (n=1), French Guyana (n=1), Uruguay (n=2), Belize (n=1), Curacao (n=1) and Argentina (n=1). Most of the studies were conducted in the years 2016, 2017 and 2018.

**Figure 1.**
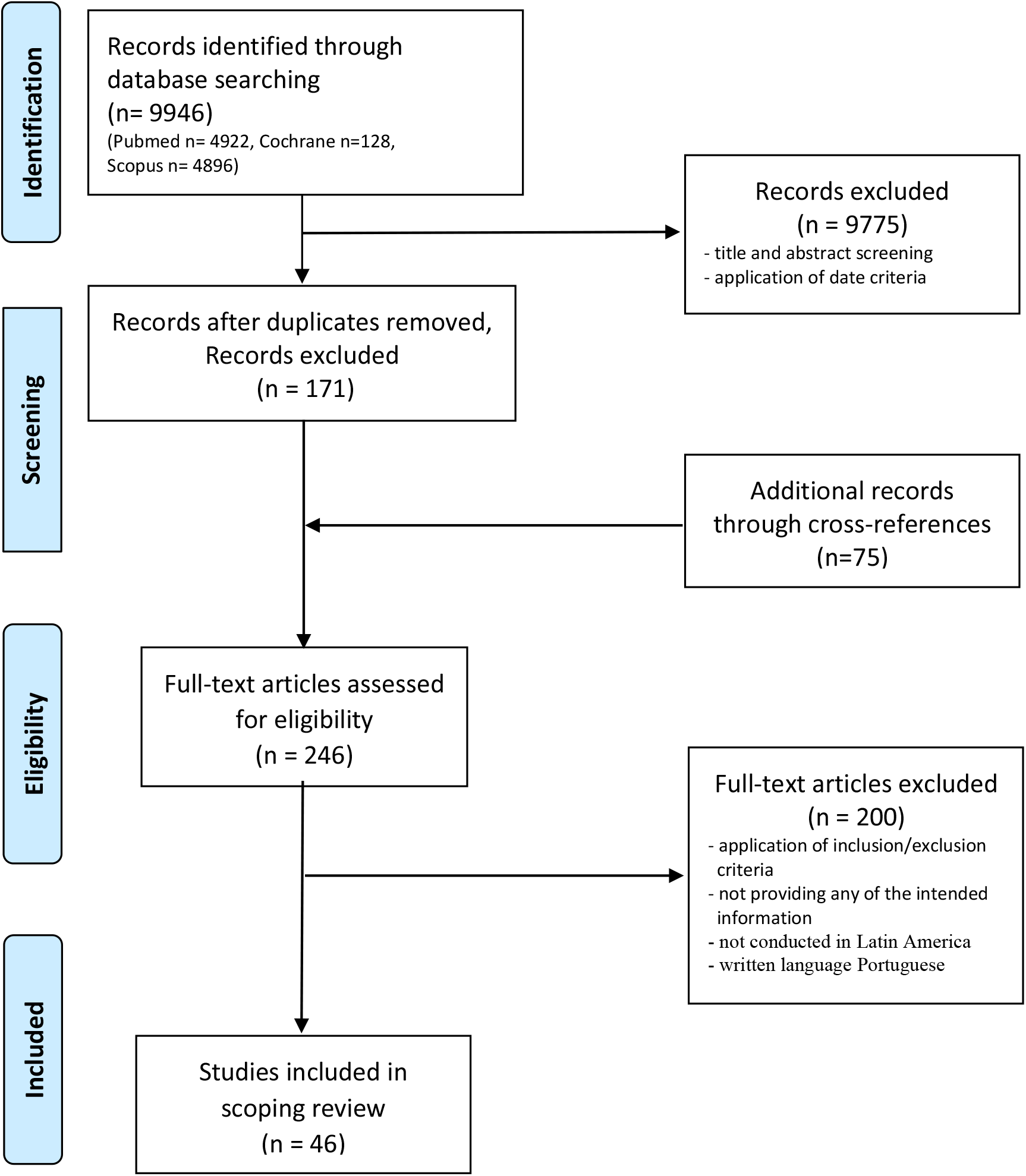
PRISMA Flow diagram of Selection Process of the included and excluded studies.

### Description of the interventions

Some types of programmes have been implemented to control the mosquito borne diseases Zika, Dengue and Chikungunya through vector management. **[48-50, 52, 53, 57, 59, 62-69]** Some studies focused on awareness healthcare campaigns about the sexual transmission of Zika virus and contraception programmes. **[8, 32-37, 54, 75]** Others discussed a family support approach and provided information about family support services. **[29-31, 61, 74]** Many publications assessed the level of knowledge, healthcare barriers and sexual health care through interviews, focus groups and surveys. **[8, 21-24, 26, 27, 36-38, 47, 52-54, 56, 60, 63, 70]** The implemented public health projects can be summarized and categorized into four broad key topics: 1) Mosquito vector control initiatives, 2) Contraception and reproductive healthcare, 3) Family support programmes and 4) Assessment of knowledge, perception and barriers. Almost all studies reported intervention effectiveness and improvement or changes in behaviour.

### Mosquito vector control

Among the studies, which reported mosquito vector control initiatives, several publications reported community engagement projects of mosquito vector control and described the active involvement of communities into the planning, developing and implementation process of an initiative. **[48, 50, 57, 59, 62, 64-66]**. A community-based participatory approach was provided among a community in the Amazon basin of Peru and at the same time among a community in Thailand. The aim of the study was the development, design and placement of a lethal oviposition trap as habitat for Aedes species. The feedback of both communities in different parts of the world was very similar and the paper highlights the community-based participatory approach for long-term sustainability. **[59]** Another community participatory approach towards mosquito vector control took place in Mexico and Nicaragua. Several studies described a larger scaled pesticide-free mosquito vector control initiative, called the “Camino Verde” (“The Green Way”) trial. **[64-66]** The Camino Verde trial used elements of the SEPA (“socialising evidence for participatory action”) process, which is an evidence-based approach to community mobilisation and participation. It consists of collaborating with communities during different phases of the process by seeking dialogue, joint discussions and solutions with community members in context of their living realities. One of the included articles reported a community participation intervention regarding vector control in Guatemala, which built on former initiatives against Dengue epidemics in Mexico and the West Nile Virus vector in Canada. **[50]** A collaboration across different sectors including academic researchers, local health authorities of the national programme on vector-borne transmitted diseases by the Ministry of Public Health and Social Assistance of Guatemala, international contributors and community members could be achieved.

Furthermore, a study of pre-post design took place from January to August 2019 and described an educational intervention on schoolchildrens’
s knowledge and actions concerning vector-borne diseases in Arequipa, Peru. **[71]** According to the results, health education in schools would be very valuable in combating future epidemic outbreaks. Apart from this project, several articles emphasized the importance of involving school children. **[58, 66, 68, 71]** The Puerto Rico Department of Health together with the US Centers for Disease Control and Prevention examined the behavioural change after a four-component intervention. **[51]** The initiative took place over a period of one year from July 2016 to June 2017 and consisted of an education program, the “Women, Infants, and Children Program Zika Orientation”, the distribution of a Zika prevention kit, a communication campaign called “Deten el Zika” (“This Is How We Stop Zika”) through media like television, radio and social media platforms, and free home mosquito spraying. The study pointed out that interpersonal communication and social context factors might have major influence on the acceptance of preventive behaviour.

Moreover, two systematic reviews performed in Argentina and Mexico examined efficacy of vector control interventions and health outcomes, outcomes of larval indices (HI, CI, and BI) after chemical and biological vector control strategies compared to community engagement strategies **[67, 68]**. Community mobilization was realised as most successful to reduce entomological indices and recommended especially to governments that use chemical vector control strategies only.

### Contraception and reproductive healthcare

As the Zika virus might be transmitted through sexual intercourse it is important to protect vulnerable population groups through comprehensive healthcare services and raise awareness about modes of transmission of the virus. Therefore, the exploration of knowledge and barriers to contraception formed an essential part of the Zika outbreak response. **[8, 32-37, 54, 75]** In 2016 a qualitative assessment was performed in Puerto Rico to examine the access to reproductive healthcare, to raise awareness about contraception during the Zika virus outbreak, attitudes towards contraception, and to find adequate risk communication strategies. **[32]** Another publication described the “Zika contraception access Network” (Z-CAN) program, which was installed as short-term response to the Zika virus epidemic to improve health outcome through increased use of contraception and subsequently less unintended pregnancies. **[33]** The program aimed to facilitate access to reproductive healthcare by reducing costs and barriers, increase knowledge and awareness in women, reinforce infrastructures, and try to ensure sustainability of services for the future. To achieve these objectives, Z-CAN built on strong intersectoral collaboration between stakeholders in the healthcare system and NGOs in Puerto Rico. Another assessment of the included studies aimed to gain information about experiences regarding pregnancy management related to Zika virus. **[34]** The Centers for Disease Control and Prevention (CDC) and The Puerto Rico Department of Health (PRDH) evaluated data of the Pregnancy Risk Assessment Monitoring System Zika Postpartum Emergency Response (PRAMS-ZPER) survey, which gathered data of women who had a live birth in the period from August to December 2016. The PRDH instructed health care providers to point out the possibility of testing for Zika virus at the beginning of antenatal healthcare to all pregnant women with or without symptoms living in areas with current Zika transmission. Furthermore, a research team conducted a study to examine knowledge and Zika virus-related attitudes among adults during an exploration concerning HIV and sexuality in the Amazon and Andes region of Ecuador. **[37]** The investigations showed an increased risk for pregnancies among adolescents and indicated the need for further sexual health education and programmes.

### Family support programmes

Following family planning services especially in context of post-Zika outbreak, a few studies discussed the need and importance of family support initiatives. **[29-31, 61]** A community-based parent group intervention “Juntos” for children with confirmed Congenital Zika Syndrome in Rio de Janeiro and El Salvador, particularly investigating fathers’
s engagement in the role as caregiver, took place from August 2017 until May 2018. **[29]** Apart from social stigma and financial challenges, this publication mentioned the increased risk of fathers abandoning their families once they receive the diagnosis of an infant’s neurologic disability. Besides, a group of researchers started an investigation about the practical and societal relevancy and utility of a family support program regarding the neurological consequences for children related to Zika virus infection. **[30]** Apart from the financial burden, barriers like distrust with healthcare personnel, communication problems, misunderstandings and stigmatization are further challenges for families. Following this approach, one publication emphasized the need for more psychosocial support and adapted healthcare regarding living conditions of families with children suffering from severe neurologic disorders to reduce vulnerability as well. **[61]** The study pointed out that especially mothers suffer from social and financial pressure, because they are perceived as the main caring person and very often stop working to be available for the child.

### Assessment of knowledge, perception and barriers

Many publications did not describe active community engagement processes but gave an insight into attitudes towards Zika virus, personal protection measures and assessed understanding and acceptance among vulnerable population groups. **[21-24, 26, 27, 38, 47, 52-54, 56, 60, 63, 70]** Perspectives, expectations and experiences of pregnant women in healthcare service access, who had been diagnosed with Zika virus during their pregnancy and given birth to a baby with microcephaly, were examined in the Zika endemic area of Villavicencio in Colombia. **[26]** The study recommended to improve sexual and reproductive health education for adolescent women or women of childbearing age as well as for their male partners during prenatal follow-up consultations. Following this, a further study discussed the barriers to health care access especially with respect to sexual healthcare services for adolescent women and young women of reproductive age living in three Zika endemic towns in the northeast of Brazil. **[23]** This publication recommended culturally appropriate information campaigns in schools or during prenatal care about pregnancy, contraception, reproductive and sexual health care; family planning services should be extended in this area as well. A study conducted in the mid-west area of Brazil assessed the knowledge, perceptions, and self-care actions of women with Zika virus infection during their pregnancy. **[54]** The results demonstrated that women obtained very little information from basic healthcare services and that there exists a deficit in prenatal care and aftercare by professionals due to lack of medical and public health services guidance. The study stated that not many publications exist in Portuguese language so that healthcare providers could not improve themselves in the provision of prenatal healthcare in accordance with current regulations.

In the Dominican Republic, a study team aimed to find out about information sources, awareness of symptoms, consequences, reproductive healthcare and prevention methods of Zika virus infection. **[21]** Differences between urban and rural communities could be demonstrated, general knowledge was very low and community residents were not aware about sexual transmission of the Zika virus. In addition, a paper-based survey took place involving 44 healthcare professionals at four clinics and 348 patients to assess level of knowledge, perceptions and prevention behaviour among the Zika virus epidemic in Roatán, Honduras. **[70]** It pointed to the importance of fostering healthcare education and the need for improvement of public messaging in risk areas.

## Discussion

This review assessed a wide range of qualitative research during the Zika virus epidemic in Latin America and the Caribbean. According to the “UNICEF minimal quality standards and indicators for community engagement” this process can roughly be divided into the four sections community engagement, implementation, coordination and integration and operations. Our findings showed several projects or studies in accordance with these criteria. **[29, 48, 50, 57, 59, 64-68, 71-73]** Some of the publications did not report engagement of community members in all processes of an intervention but involved them at least in some parts of the projects **[30-33, 37, 47, 49, 51, 60, 62, 75]**. A lot of studies described qualitative research through semi-structured interviews, focus group discussions and approaching vulnerable populations during prenatal care and home visits or via online surveys. Experiences, knowledge, prevention measures and perceptions towards Zika virus infection were assessed, but without any engagement in planning and implementation processes, decision making or empowerment activities. **[8, 21-24, 26, 28, 34-36, 38, 52-54, 56, 58, 60, 61, 63, 69, 70, 74]** However, most of the studies aimed to encourage and support behaviour change by addressing the preventive actions, that have been taken or should been taken to avoid infections and raised awareness about the risks of a Zika virus infection.

### Engagement of stakeholders

In order to successfully start and implement a public health project, it is essential to cooperate with local government authorities or stakeholders in the healthcare system to achieve effective decision making, build trust, have feedback loops, reach cost-reduction and public and social accountability. **[42, 43]** Governments have responsibilities towards their citizens to provide social services and manage environmental issues. Besides, building on local capacities is an important issue not only to save costs but to understand the circumstances, have an impact on already provided services on government level, community and individual level and work on expansion of these existing services and programmes. One study described the quick implementation of a contraception network between health professionals in Puerto Rico, who were trained in provision of free contraception methods and knowledge about Zika and could improve local healthcare services and access but demonstrated future challenges in cost-reduction or coverage of long-acting reversive contraception methods. **[33]** A greater capacity of the health system in Puerto Rico could be reached in this case but an expansion of health insurance coverage to provide more expensive services should be discussed on government and international level, as money of private donors was included but cannot always be guaranteed. Some publications offered information about existing mosquito control programmes by the government or Ministries of Health and were enhanced through the reported public health intervention without any donations. **[32, 48, 50, 56, 62, 66, 72, 73]** Altogether, the results pointed out and confirmed that engagement of stakeholders, intersectoral cooperation and building on local capacities can foster the implementation of public health interventions and strengthen the expansion of existing services for community benefits.

### Implementation and participation

A couple of studies described a detailed participatory planning process involving community leaders and residents into the public health intervention. **[29, 48, 50, 57, 59, 64-66, 68, 71-73]** For instance, community members were engaged in design, implementation and evaluation of the development of a mosquito ovitrap to practice vector control. **[59]** The study was conducted in parallel in a community in Peru and in Thailand. Findings showed a similar result in distant parts of the world with different cultural circumstances, which means that following a good structured engagement process leads to efficient integration of novel strategies on the part of target populations and strengthens long-term commitment to apply the new method. Numerous studies provided evidence of social interaction to be an effective way of learning and integrating new behaviours. **[51, 57, 59, 62, 64-66, 69, 72, 73]** In context of health education projects, community residents learned about mosquito biology, elimination of breeding sites and modes of transmission and were engaged in social activities. Periodic meetings, community health forums led by group moderators, town hall meetings and neighbourhood gatherings provided a place to share information, communicate, discuss, give feedback, exchange experiences and socialize. **[33, 50, 57, 62, 65, 72, 73]**

Furthermore, many of the papers described the engagement of so-called key persons in a community as crucial. **[24, 32, 47-50, 57, 58, 64-66, 67, 72, 75]** As the UNICEF recommends, community leaders, community mobilizers and facilitators should be selected to build trust, to accompany or guide and monitor the initiatives. **[14, 43]** They should reach agreement on activities and better comprehension of the addressed core issues of the projects within the community. Usually, facilitators belong to the target community, know the living conditions and can encourage conversations with residents. A community participatory process was conducted in Mexico and Nicaragua using an evidence-based community mobilization tool called “Socialisation of Evidence for Participatory Action” (SEPA) **[64-66]** Further campaigns we found were “The Deten el Zika” campaign (“Stop Zika”), which included the delivery of mosquito repellents and contraceptives, HIV and ZIKV information sheets and donation to a public health clinic. **[37, 51]**. The “Ante la duda, pregunta” (“When in doubt, ask”) campaign focused on conversations between healthcare providers and women concerning Zika. Another recommended approach to avoid a Zika virus infection is the establishment of educational campaigns in school. Though, only a few studies emphasized the importance of educational initiatives in schools and the involvement of teachers, who can offer interaction and creative participation and provide information and instructions in an accurate manner. **[58, 66, 68, 71]**

Another point of engaging communities into public health response to the Zika virus epidemic was the consideration of the time after an outbreak. Families or parents with children of Congenital Zika Syndrome must deal with a higher burden of psychosocial and financial stress. Parent groups can play a supportive role in avoiding social exclusion due to stigma, provide a space for exchanging experiences and discussions with other parents of affected children, learn to improve care and help families and their children to be active members of society. **[29-31, 61]** Findings demonstrated that collective community actions can raise social awareness and neighbourhood responsibility, therefore strengthen the realization and success of an intervention and enable many community members in participating and thus improving common health-outcome.

### Communication

Initiatives should be communicated clearly to community members, which might be only possible in local language to avoid misunderstandings, and therefore it is necessary to be familiar with the local dialect or involve resident personnel. A few studies mentioned that the use of a complicated, medical language or unkind behaviour in health facilities resulted in misconceptions and fear of questioning, especially in groups of people with lower education levels. **[23, 30, 35, 48, 51, 54, 62, 73]** Apart from clinic services or health professionals and providers, most popular sources of information and delivering messages were television, radio, word of mouth by family and friends, and social media **[22, 32, 37, 38, 51, 53, 54, 56-58, 60, 62]** Regarding the risks of a Zika virus infection, some studies reported scepticism towards the government and the media coverage about prevention measures because often, none of the immediate circle of acquaintances suffered from a confirmed Zika virus infection. Risk communication was perceived as way of spreading fear among populations by the government to raise sales and earnings of mosquito repellents or as invention to have an impact on fertility behaviour. **[23, 26, 30, 32, 47, 51, 54, 57, 69]** Some publications noted insufficient communication about postponement of parenthood during the Zika outbreak. Care-seeking women were given advice about mosquito prevention measures but did not receive information about the possibility of sexual transmission of Zika virus during pregnancy. **[8, 22, 28, 38, 53, 56, 63, 70]** This might be due to social taboos and religious reasons as Latin American countries are mostly catholic, and shame feelings around sexuality and difficulty in talking about sexually transmitted diseases for both, healthcare providers and community residents exist. Therefore, adequate communication of Zika virus risks should take place in healthcare services during antenatal check-ups and involve the male partners into comprehensive explanatory dialogues to get rid of false perceptions. Governments and other stakeholders must make sure to provide updated health protocols to all primary healthcare centres and clinics, train healthcare personnel and inform the public in language that is understood by all groups.

Few publications discussed laboratory testing during pregnancy, which is recommended for pregnant women living in Zika endemic areas but often not feasible in practice due to costs or lack of laboratory resources. Though some women paid services privately they still faced long waiting periods or delayed test results. Moreover, continuity of care could not always be provided because of shortfalls of specialised doctors and therefore limited possibility to attend medical appointments. **[26]** Hence, our findings point to a need for improvement of access to laboratory testing for symptomatic and asymptomatic women during their prenatal care visits. Apart from establishing more health education campaigns to increase knowledge, avoid misconceptions and misguided assumptions within a community, a better collaboration between stakeholders might lead to a higher service availability and thus can improve quality of life for all population groups.

### Limitations

The scoping review assessed a large number of publications all over Latin America and the Caribbean to gain an insight about implemented intervention projects related to the Zika virus outbreak. There might be limitations in scope due to the uneven geographical distribution of approaches as most of the studies were conducted in Brazil or Puerto Rico. Furthermore, the authors have good knowledge in English and Spanish but capacity to identify, include and translate articles in Portuguese was limited. However, during the literature search the minority of papers appeared to be published in Portuguese and as only three Portuguese papers were considered for a closer selection this might be of little consequence for getting an overview on the topic.

## Conclusions

Findings highlight free decision making processes, social activities and educational entertainment as most productive and successful ways of implementing a community engagement project in context of the Zika virus epidemic. Several educational community participation programmes resulted in reduction of Aedes aegypti populations due to collective elimination of breeding sites and increased personal protection measures to avoid mosquito bites or sexual transmission. Awareness and knowledge of transmission routes and consequences of a Zika virus infection could be raised in vulnerable population groups.

Collaborative partnerships between study teams, local health sector and local government agencies are an important component for financial support and health improvement in general and concerning the Zika virus outbreak as well. Not imposing duties but involvement of communities into planning, decision and operational processes leads to high acceptance of projects and is most beneficial for sustainability. Learning by doing and communication at eye-level ensured empowerment of community residents and fostered a sense of community.

## Data Availability

All data generated or analysed during this study are included in this published article.

## Funding

No funding to declare

### Ethics and dissemination

Ethical approval is not required because this study does not involve collection of primary data.

### Availability of data and materials

All data generated or analysed during this study are included in this published article.

### Contributors

SN conceptualised the research, prepared the draft article and wrote the final manuscript; RK conceptualised and supervised the research and reviewed the different versions of the manuscript

### Patient consent for publication

Not required

### Provenance and peer review

Not commissioned, externally peer reviewed

### Open access

