## Supplemental Table 1 for "Community participation and community engagement in the response to the Zika-Virus-Outbreak in Latin America 2015-2019 – A Scoping Review"

1.1. **Charting map of included studies**

**Table 1**  Charting Map of included studies in the Scoping Review

| **Author** | **Country, Year of Publication** | **Study design, Year of Conducting** | **Aim of the study** | **Components of intervention** | **Knowledge, Perception, Barriers and Main Findings** | **Challenges faced, lessons learnt and recommendations** |
| --- | --- | --- | --- | --- | --- | --- |
| August EM et al. [32] | Puerto Rico  2019 | qualitative study  8-9/2016 | -) attitude towards contraception  -) barriers and use of contraception  -) communication strategies about the availability through Z-CAN | -) women/men aged 18-49  -) 10 focus groups  with 5-8 participants  -) contraception awareness campaigns: “Ante la duda”, “Yo tengo la  voz”, “Ya tu sabes” | a) knowledge: internet, social media, friends, family, physician; oral contraceptive pills, condoms, fertility-awareness methods  b) Perception: information scepticism: fear, increase repellent sales, influence reproductive behaviour  c) barriers: costs  -) limited awareness because Zika mostly asymptomatic  -) motivator for contraceptive use: costs linked to unintended pregnancy; preferred concept Ante La Duda: dialogue between woman and hcp, less focus on Zika virus) | -) more information about free contraception services (location, availability, costs)  -) more information about side effects/effectiveness of contraception methods  -) trustworthy contact persons important  -) digital strategies useful |
| Lathrop E. et al.  [33] | Puerto Rico 2018 | cohort study 4/2016-12/2017 | -) increase access to contraception/reduce barriers (cost, service points, lack of providers) for pregnancy prevention in context of Zika  -) prevention of unwanted pregnancy to decrease adverse pregnancy related to Zika virus  -) raise awareness of contraception methods as prevention | -) short-term response  -) “Zika Contraception Access Network” program: network of hcp (doctors, clinic staff) trained in contraceptive counselling and same-day provision  -) recruitment participants:  1-day training, through hcp, word of mouth, health education campaigns, involving community engagement activities, internet campaigns  -) satisfaction survey | -) before Z-CAN: no use/ineffective contraceptive methods (condoms, sponge, withdrawal, spermicide, fertility awareness methods); rare male sterilisations, rare provision of contraceptive implants or levonorgestrel-releasing intrauterine device in clinics  -) barriers: limited contraceptive access by policy, finances, logistic, few patient education  -) 8/2017: 21 124 women`s data available with Z-CAN  -) 20 110 received same-day provision of reversible contraceptive method (68% LARC method, 25% oral contraceptive pills/hormonal contraception, 5% no method)  -) 3068 participated in satisfaction survey and were satisfied  -) quick capacity building with purposive training sessions possible | -) partnerships between programme teams and stakeholders fosters programme  -) improved access to contraception decreases unwanted/adverse pregnancies  -) capacity expansion of health-care system  -) cost challenges (continued availability of LARC – cost-sharing in health insurance=  -) adaption of Z-CAN in other settings |
| Leontsini E. et al.  [47] | Guatemala  2020 | qualitative study  7-8/2017 | -) exploration of cultural salience, effectiveness, collective  efﬁcacy, feasibility of Zika prevention actions and recommendations  -) Data for improvement of social and behaviour change communication (SBCC) efforts | -) 2 study sites  -) (pregnant) women, aged 18–30 and partners  -) 70 participants  -) 12 focus groups,  -) community outreach  worker/fg moderator  -) 19 task cards; “Strategic Communication for Zika Prevention: A Framework for Local Adaptation” by USAID + UNICEF to guide country-level communication strategies” | a) knowledge: mosquitoes come with rainwater; no awareness that Ae.ae. attaches eggs to inner walls of water containers (not directly in water) no distinction between mosquito types (Ae. aegypti = day biter)  b) perception: less concern (mild symptoms, reduced media coverage in contrast to recent Chikungunya); fear of consequences for babies; “invention of health centre”  c) prevention: >32 Zika preventive actions, common mosquito control: bed net use, elimination of rainwater containers, repellents, general house cleaning and surroundings, use of bleach to disinfect water but no application to destroy mosquito eggs; gender roles interfere with condom use to prevent sexual transmission of Zika | -) more intersectoral collaboration (waste services)  -) Increase collective self-efﬁcacy for long-term preventive behaviour  -) Increase self-efﬁcacy for condom use among young adults without family plans, promotion of antenatal care, family planning counselling |
| Juarbe-Rey D. et al. [57] | Puerto Rico  2018 | qualitative (2 months period) & quantitative,  12-month period | -) impact of risk communication strategies for prevention and control of Zika virus  -) knowledge, attitudes, behaviours regarding Zika  -) increase awareness,  engagement and mobilization  -) cooperation: Manuel A. Perez housing project, Graduate School of Public Health, Medical Sciences Campus, University of Puerto Rico | -) women of reproductive age, mothers, community and sport leaders, students  -) risk communication initiative, community-  based participatory process “Unidos contra el Zika”  -) 335 participants  -) focus groups  -) community advisory  board  -) face-to-face interviews  -) awareness health fair  -) educational weekly meetings, theatre, exchange with community  -) follow-up interviews | a) knowledge: tv, community activities, handouts, internet, little via health providers  b) Perception: sceptics because of previous experiences with academic researchers “helicopter research” (no benefits for community members, own advancement for researchers)  c) prevention: Mosquito control activities (emptying and cleaning containers, remove standing water), 60% feel personal responsibility  3 risk communication strategies:  -) Zika awareness health fair (140 participants)  -) health education through theatre (130 participants)  -) community forums (75 participants)  -) adoption of Zika prevention and control practices (site reduction), high susceptibility among community members  -) pre-/post-survey: responsibility increased 60% to 83% (behaviour change) | -) establishing community-academic partnership is time-intensive  -) building trust through frequent meetings  -) development of culturally appropriate strategies  -) health information through internet, Facebook for risk communication  -) usefulness across different populations would be interesting (study conducted in a low-income community) |
| Parker C. et al.  [48] | Honduras  2019 | pilot study  Dec. 2018 | -) create community-based vector control program for container breeding mosquitoes  -) Universidad Nacional Autónoma de Honduras  -) Ministry of Health carries out mosquito control in response to an epidemic also | -) underserved community  -) participants: community leaders, stakeholders (university/students, doctors, nurses, engineers, government oﬃcials)  -) workshop on mosquito biology, ecology, control (lectures, hands-on activities)  -) community outreach / community-based campaign  -) educational material distribution  -) identify larval habitats, container elimination  -) pre-/post-survey | a) knowledge: basics before and after training; majority had heard of Zika, dengue, or chikungunya (6% did not), impact of awareness of vector control activities on mosquito density, primary education more containers  b) prevention: 13% covered containers to prevent oviposition (6% of participants more containers), common: ﬂowerpots, garbage, toys, water storage containers, buckets, water-holding plants, water drainage, trash bins  -) Aegypti and Ae. albopictus, short ﬂight range (200m, max 800m), neighbours  -) community leaders and stakeholders able to spread knowledge after training; identify and target containers  -) successful control of container mosquitoes depends on participation of community leaders, stakeholders and community itself  -) transmission risk: combination of social, ecological, and climatic conditions | -) engaging community in vector control enables residents  -) engaging stakeholders facilitates IVM program  -) more intervention time  -) improvements in workshops and expansion of program to other territories (provide workshops in community´s language; result varies (region; dry/rainy)  -) more vector control campaigns & frame-work for training in engaging  -) poverty and vector-borne diseases associated |
| Duman-Scheel M. et al. [69] | Belize  2018 | internet-based assessment survey,  3-4/2017 | -) importance of mosquito control to stakeholders in tourism industry, Belize  -) current mosquito  control practices  of stakeholders  -) perceptions and attitudes towards new mosquito control technologies  -) Indiana University Office of Research Compliance | -) 168 participants  -) 64 establishments (hotel/resort, tour operator, restaurant, other)  -) survey invitations via email (combination of 40 five-point scale items, fill-ins, open-ended writing prompts)  -) protect citizens and visitors (weekly surveillance, monthly health fairs by Belize MoH vector program) | a) knowledge: good knowledge of mosquito vector, mosquito control on properties (breeding sites, insecticides)  b) perception: concern for unborn, over-sensationalizing by media therefore negative impact on livelihoods and business; media coverage educates people  c) prevention: correlation of larvicide use with willingness to use insecticides, belief water treatment decreases mosquito densities and disease transmission; 91% use insecticides  -) vector control important for tourist-based industry  -) minimal impact on environment or health wanted  -) product purchase choices: safety of products (humans, animals, plants) product effectiveness; lack of interest correlates with lack of willingness | -) importance of community engagement  -) new mosquito control technologies: eco-friendliness and sustainability; problem: insecticide resistance, effects of pesticides on nontarget species  -) weighing benefits and costs of media coverage about infectious health threat |
| Castro, M. et al.  [72] | Cuba  2017 | perspective Essay,  11/2016 | assessment of the Cuban response to Zika | key strategies:  -) vector control  -) surveillance of  cases and symptoms  -) training of health professionals  -) risk communication  (mass media, community health forums)  -) mobilization | -) universal health-care system  -) early declaration of national alert for Zika 12/2015  -) early and successful response to Zika based on dengue prevention/control programs (experimental studies, randomized control trials, process evaluations and cost analyses since 2000)  -) political will  -) active community engagement and participation; establishing teamwork mechanisms  -) promotion of preventive behaviours through mass media communication with messages  -) entry of Zika into Cuba impeded through integration of risk communication strategies | -) model for quick mobilization and intersectoral collaboration  -) application of dengue strategies (CE) to Zika outbreak  -) need for community capacity-building |
| Gorry C.  [73] | Cuba  2016 | descriptive Essay,  2016 | description of  measures taken in Cuba in response to Zika | -) national 11-point Zika action plan “All Hands on Deck” strategy  -) based on PAHO guidelines  -) cooperation with authorities  -) vector-control  measures  -) media campaign  -) neighbourhood meetings  -) protocols about surveillance  -) treatment  -) monitoring of pregnant women for medical staff | -) 2/2016 Zika Action Plan: detect, prevent, respond;  -) Daily health ministry meetings collected data  -) Weekly meetings with all authorities (MoH, Tourism, Industry, Economy, Planning, Civil Defence)  -) vector-control: participation local health authorities and communities and of 9000 soldiers in fumigation of homes, ofﬁces, schools, garbage pickup, repairs of public infrastructure (open sewers, water leaks)  -) education and media campaign: elimination of breeding sites, fumigation, mosquito repellent, protective clothing etc  -) neighbourhood meetings with doctors (symptoms, warning signs, door-to-door visits by medical students)  -) meetings with homeowners who rent to travellers  -) update of international health regulations in Cuba (cooperation programs instead of closing borders/lock down), identify travellers with fever (thermal imaging scanners at airports, ports, marinas), 10 days medical observation before returning to Cuba, hospitalization in case of fever  -) standard antenatal check-ups regarding Zika: national protocols (surveillance, treatment, monitoring of pregnant women) | WHO/PAHO lessons learnt from Ebola to apply on Zika:  -) standard protocols, accurate, up-to-date, understandable information for health care providers and public  -) political will, global/local community cooperation, funding, implementation of standard protocols for preventing, controlling, diagnosing and treating (re)-emerging diseases  -) national dengue diagnostic and surveillance network since the 70s |
| Barrera R. et.al  [49] | Puerto Rico  2019 | qualitative study  11/2016-8/2017 | -) integrated vector  management  -) reduction of transmission by detection of viruses in Ae. aegypti population  -) percentage of vector control coverage for effective reduction  -) assessment if mass trapping possible to use in medium size cities | -) Caguas City  -) 6 focus groups  -) IVM intervention: 1) community awareness, education, 2) source reduction and larvicide with mass mosquito trapping trough ovitraps, 3) monitoring of rainfall, temperature, humidity, assuming mosquito life span of 3 weeks  4) analysis of female Ae. aegypti adults for foreign RNA (Dengue, Zika, Chikungunya) | -) initial goal: treatment of at least 80% buildings  -) installation of 3 mosquito traps per home  -) containers: water meters, trash cans, cavities, bromeliads, plastic buckets, flowerpots, barrels, lids on top of buckets or barrels  -) community leaders as effective facilitators  -) barriers: residents’ absenteeism (revisits up to 3 times), local circumstances (composition of buildings), more training for field personnel (use of maps, electronic devices for data)  -) no mosquito population reduction with treatment coverage/ percentage of buildings below 20%  -) control Ae. aegypti through mass trapping adult female mosquitoes, AGO traps attract and capture gravid/female mosquitoes | -) mosquito density changed with intervention  -) effective IVM with mass trapping  -) use in medium sized city possible  -) variations between  clusters (composition of buildings, residents available or willing to participate) |
| D’Angelo, D.V. et al. [34] | Puerto Rico  2016 | descriptive survey  1/2016 – 3/2017 | -) experiences regarding prevention and diagnosis of Zika virus infection during pregnancy in PR  -) analysis of data from the Pregnancy Risk Assessment Monitoring System Zika Postpartum Emergency Response (PRAMS-ZPER) survey (CDC, Puerto Rico Department of Health) | -) 2364 women (data of 3,300 pregnant women  with laboratory evidence of possible Zika virus infection 1/2016 to 3/2017)  -) 36 hospitals  -) approach of women with life-birth during  hospital stay  -) raising awareness  about virus and prevention ways  -) promotion of prevention of Zika transmission | a) knowledge: familiar to women (similar guidance in campaigns related to dengue and chikungunya), 94.3% information by health care provider about ZIKV infection during pregnancy (transmission, prevention, use of condoms); 76.9% tested for Zika virus by hcp during 1st or 2nd trimester  b) perception: worries about infection during pregnancy (93%), about possibility of birth defects; for 70% hcp best information source about ZIKV  c) prevention: 38.5% abstain from sex/ use condoms during pregnancy; common reasons for rare condoms use: belief partner does not have Zika virus/ condoms unnecessary during pregnancy; 98.1% mosquito bite protection (closed doors and windows, removing standing water, larvicide, mosquito net); less use of long-sleeves, repellents | -) reinforcement during prenatal care visits and public communication campaigns  -) gaps in use of preventive measures – more prevention messages, safety of repellent use, sexual abstinence or condom use during pregnancy  -) increase testing for Zika virus during pregnancy  -) third trimester testing added by PRDH |
| Nelson E.J. et al. [21] | Dominican Republic  2019 | qualitative study,  10-24/5/2017 | knowledge of symptoms, health effects, prevention practices related to Zika virus in communities, northern coast of the Dominican Republic | -) 75 participants  >18 years  -) interview invitation while attending free medical clinics  -) 4 domains: knowledge, attitudes, practices and respondent perceptions  of ZV  -) WHO KAP survey  tool, reduced from 155 to 41 questions | a) knowledge: differences between rural and urban areas, low knowledge about basic risks (vulnerable groups, transmission, prevention, symptoms) and sexual transmission of Zika, consequences of infection during pregnancy in study sample, only 5 women (10.4%) aware about neurologic consequences, 40% mosquitoes or sexual transmission, for 51% Zika important concern  b) perceptions/concerns: becoming sick (40%), babies being born with disabilities (19%), fear Zika is contagious (9%)  c) prevention: cleaning household environment (33%), mosquito nets and repellent (13%); 55% no prevention actions (interviews conducted one year after CDC statement Zika related to microcephaly)  -) Low education level/incomes in rural communities | -) more accessible information and educational campaigns about Zika-related health risks, particularly for pregnant women / sex tourism  -) few trained medical doctors and accessibility to specialists  -) Zika-related information from medical professionals highly accepted |
| Ulibarri G. et al.  [50] | Guatemala  2016 | qualitative study,  10 months period, 2015 | -) effectiveness of  integrated health intervention in risk populations of Zika virus transmission  -) adoption of administration, care and sustainability, empowerment of community by trained health providers  -) collaboration MoH Guatemala vector control programme, National Institute of Public Health Mexico and community | -) 16 focus groups/8 participants per group  -) 3-component  integrated intervention over 10 months  -) web-based vector control training for health workers  -) use of ovillantas (low-cost ecological mosquito control)  -) pre/post-interviews | a) knowledge: very low among community members and health workers  b) perception: mosquitoes breed only in natural ponds, cleaning of house/garden are women´s tasks, MoH’s services not efficient – preference for self-medication (local medicinal plants); Low regard for advantages of community participation, low acceptance of new mosquito control methods & prevention strategies  c) barriers: few information provided about cause of illness and consequences; Dependency on public services (cleanliness on streets/on public antimalarial control); lack of internet access  -) after intervention: increased knowledge, interest,  participation in community mosquito control and trapping, acceptance of ovillantas; cleaning houses and gardens | -) intervention beneficial, willingness to participate  -) fostering collaboration and  community participation recommended  -) difficulties to access computer equipment, lack of training in computer use, no or slow internet at home/in the region |
| Brisset D. et al. [70] | Honduras 2018 | paper-based survey  (March–May 2017) | -) evaluation of ZIKV knowledge, attitudes,  and preventive practices  -) outcomes 6 months after public health educational campaign, attention to sexual health  -) former study in Roatán 2016: blood samples and clinical information from 183 patients with suspected dengue virus infection; 79 patients Zika positive | -) 348 residents > age 18  -) 44 Health Care Providers at hospital  -) 4 clinics in Roatán  (public, private)  -) invitation during medical health outreach programme or community service activities  -) WHO “Knowledge, Attitudes and Practice” questionnaire  -) survey of 37 questions | a) knowledge: difference between HCPs and residents: majority knew about mosquito transmission, low knowledge about sexual transmission; not well informed about risks, complications and prevention of ZIKV infection, <50% of residents informed by HCPs regarding prevention actions.  b) prevention: 34-68% elimination of standing water; protective clothes, use of mosquito net, low sexual abstinence, low use of condoms though available, >50% believe women should avoid pregnancy if ZIKV exposure  c) perception: majority against abortion, for 54% HCPs and for 68.4% residents severe health concern  -) survey adapted from WHO KAP survey: Zika virus disease and potential complications Resource Pack”  -) rare knowledge of sexual transmission hinders elimination | -) variety in knowledge  -) more and better educational initiatives about transmission  -) Improvement of public health messaging in ZIKV-exposed areas  -) Focus on mosquito control  -) Improvement in preparedness during future outbreaks |
| Elsinga J., et al.  [58] | Curacao  2017 | cross-sectional mixed methods study,  6-7/2015 | -) investigation of success of achieved mosquito control  -) exploration of media role, coverage and improving messages to public | -) lab. confirmation patients of chikungunya infection 2014-2015, sample from general practitioners across the country  -) invitation + interview at homes (questionnaire)  -) recruitment: snowballing key persons, neighbourhood centres  -) 50 participants  -) focus group discussions  -) media education (tv, radio, newspaper) | a) knowledge: good  b) perception: Personal protection less effective (repellents, long sleeves); belief “healthy eating’ prevents from mosquito bites, perceived benefits but low perceived barriers to perform MBSC  c) prevention: reduce mosquito breeding sites: water source management, removing car tires in yards; low use of repellents; household with mosquito breeding sites threat for neighbourhood (short flight range)  d) barriers: difference between policy and communities’ realities  -) MBSC measures by government insufficient, engagement of key persons  -) after interventions more MBSC in community; key persons motivate; target: weekly check of house and yard for mosquito breeding sites and elimination | -) improve information access, media attention  -) network of local key persons would be good  -) enhance visibility of government’s MBSC policies  -) raise communities’ sense of responsibility  -) reduce water storage  -) more education in schools – better community knowledge  -) collaboration communities and governments essential |
| Earle-Richardso G., et al. [51] | Puerto Rico, 2018 | qualitative study  7/2016 – 6/2017 | influence of community education efforts on pregnant women’s Zika prevention behaviour during the Puerto Rico Department of Health Zika virus response 2016 | -) 1 329 pregnant women  -) monthly telephone interviews with WIC participants  -) 4 interventions to maximize self-protection behaviours:  1) PRDoH Women, Infants and Children WIC (women, infants, children) Program Zika  Orientation  2) Zika Prevention Kit Distribution  3) The Detén el Zika campaign (“This Is How We Stop Zika”)  4) Free Residential Mosquito Spraying | -) protective behaviour: 90% eliminate standing water, 44% condom use, 28% mosquito repellent use, receiving Zika prevention kit associated with larvicide and bed net use; 4% long-sleeved shirt.  Ad 1) 93% high exposure; 20–30-minute presentation on Zika to women, Ad 2) 75% ZPK distribution (repellent, condoms, mosquito bed net, larvicide); most positive influence on Zika prevention behaviour, logistical problems, Ad 3) 51%; through tv, radio, print, and social media channels, preventive behaviour on TV; greatest effect on removing standing water and repellent use; exposure grew over time, Ad 4) services enable women to handle cost and logistical barriers; received services: 34% over the whole time period, 68% for some months  -) association of exposure with implementation for each intervention  -) social context factors more important for effect on preventive behaviour than personal risk- and self-efficacy factors | -) advantages of cooperation with local partners  -) ZPK distribution effective, behavioural support  -) effects on health-related behaviour change influenced by interpersonal communication  -) findings suitable with PRAMS Zika Postpartum Emergency Response (PRAMS-ZPER) study of postpartum women |
| Heydari N., et al.  [52] | Ecuador, 2016 | qualitative design;  3-8/2015 | -) investigation of economical and socio-behavioural factors  inﬂuencing mosquito control in households of Machala  -) available mosquito control products in local markets  -) private costs on mosquito control, factors to choose mosquito control product | -) 40 households  -) semi-structured questionnaire  -) Household surveys  -) 5 variables: a) home ownership, b) prior chikungunya or dengue infection, c) stability of job d) household income below minimum wage, e) gender  -) in conjunction with a pilot ﬁeld trial of to decrease indoor density of Ae. aegypti populations | -) newness and unfamiliarity of Zika: impact on risk perception and adoption of preventative strategies  a) knowledge: good on transmission of dengue, severity and risk of dengue fever  b) perception: high regarding the severity of Zika  c) prevention: 5 different mosquito control interventions: aerosols, liquid sprays, repellents, mosquito coils, unimpregnated bed nets  d) barriers: economic limitations (effectiveness, cost); low-income communities spend >10% of family income on mosquito protective behaviour and disease prevention, limited access to public services and utilities (garbage collection, sewerage, piped water)  -) product decision: effectiveness, low cost, easy to use  -) vulnerable communities face greatest social and economic burden | -) distinction between Ae. aegypti-transmitted illnesses important for risk perception and application of appropriate vector control strategies  -) more economic research  -) minimal health risks of prevention strategies to families have greatest chance of widespread use |
| Basso C., et al. [62] | Uruguay, 2017 | cluster randomized controlled  trial.  4-11/2015 | -) scaling up process of an intervention to eliminate Aedes aegypti breeding sites  -) transdisciplinary analysis of eco-bio-social factors  -) MoH, Municipality, involvement of Consultancy and Orientation Services of the Ministry for Social Development MIDES | -) 238 participants  -) information: radio, tv, written press (time,  place, characteristics of activity), car with loudspeaker  -) 12 activities with social groups, schools, comm. Organizations  during 4 weeks  -) distribution of plastic bags for collection of water containers  -) recollection and recycling by MoH and Municipality vector control services  -) household and entomological surveys | -) monthly meeting of discussion group  -) feasible and easy application of vector control measures wanted  -) entomological survey: identify and quantify larvae/pupae; mosquito eggs laid on walls of containers near water surface, withstand desiccation embryonated for up to 1 year  -) prevention most breeding places eliminated (breeding places removed in 50% of visited households + additional plastic bags with containers);  -) barriers: financial and logistical, absence of residents during the day; political situation – municipality did not collaborate  -) key to success: Community engagement and inter-sectorial partnerships, high acceptance of intervention and willingness to participate | -) inter-institutional cooperation, action plans (environmental, bio-ecological, anthropological, logistic, communication aspects)  -) community mobilization increases removal of containers  -) targeting most productive container types for adult Ae. mosquitoes improve: efﬁciency of labour, cost reduction, elimination of adults  -) large scale intervention saves costs (training, transport) |
| Muñoz del Carpio-Toia, A., et al. [71] | Peru, 2019 | pre-post design study;  1-8/2019 | -) identification of  knowledge and practices in schoolchildren before and after an educational intervention | -) 3 schools around Arequipa  -) 300 schoolchildren between 6 and 15 years (rural, urban, urban-marginal)  -) audio-visual material: images, voices, musical background, banners,  talks, advertising spots  -) pre-/post survey of educational intervention | metaxenic diseases: dengue, yellow fever, malaria, chagas disease, leishmaniasis, bartonellosis, chikungunya and Zika  a) knowledge on diseases (global, agent, symptoms, prevention and complications); regular global knowledge in 99% of students, rare knowledge about symptoms, and 54.3% schoolchildren about the vector--) (post-intervention): level of knowledge improved in all students after educational intervention  -) cultural adaption of material  -) health education plays important role in disease prevention  -) schoolchildren empowered against diseases | -) more community mobilization to induce behaviour change, more education  -) more interaction and participation of agriculture, education, fisheries and labour sectors  -) development of educational materials for school-teachers |
| Paz-Soldan V.A., et al. [59] | Peru, (Thailand, US),  2016 | mixed methods;  2010 | -) illustration of design and placement process for an attractive lethal ovitrap to reduce vector populations  -) lessons learned in the development of the trap | -) community-based participatory approach based: perceptions + feedback, entomological findings in the lab, design + research team observations  -) interdisciplinary research team: entomologists, social scientists, industrial designers, the targeted community  -) focus group discussions  -) semi-structured interviews | -) 6 phases–process: documentation of study areas, discussions design and research team (22 potential trap designs), FG discussion (6 trap designs discussed), again FGD and testing 2 traps for 1 week, final trap design produced and shared with vector control officials  -) community-based participatory process addresses needs and concerns of community members (living situations, realities)  -) similar discussions in Peru and Thailand, despite different landscape, culture, language, type of homes  -) similar trap rankings in both countries  -) acceptable characteristics of traps: safety (little children), weather durability (rainy, windy, easy to install, stable to be used outdoor), cost, environmental impact, maintenance and aesthetics; attractive enough to own, but not desirable objects for theft; cave: potential misuse (as lamp) | -) key: structured and directed focus group guide  -) high acceptance of trap placement in homes because of the participatory process  -) focus on trap effectiveness within the community and long-term acceptability  -) discussions how to increase use, adapt to other vector-borne disease control tool development processes |
| Quintana Salced A.E., et al. [63] | Colombia, 2019 | descriptive study with a quantitative approach | -) evaluation of the social impact on the project “Mitigation of Zika virus infection" in Cartagena  -) prevention and control activities  -) national project of Compasión Internacional organization | -) 237 families, middle  age 40  -) checklist  -) house to house visits  -) survey | a) knowledge: families know about Zika virus and most important infection prevention measures, transmission by mosquito; no knowledge about treatment; recognized symptoms: 93 % fever, 70 % headache, 54 % joint pain, 28 % rash;  b) prevention: fumigation, control/elimination of standing water, environment cleaning, repellent use, insecticides, adequate clothes (long-sleeves, trousers); community education and participation effective approach to Zika virus infection  -) malformations and prevention related to sexual transmission not mentioned | -) reinforce prevention methods: use of mosquito net/fly screens on doors and windows, container covering;  -) more cooperation between national health system and locals  -) measures effective for families to participate in prevention and control |
| Smythe T., et al.  [29] | Brazil, 2019 | qualitative study;  8/2017 – 5/2018 | involvement of fathers in “Juntos”, a community-based group initiative for children with congenital Zika syndrome (CZS) and their caregivers in Brazil | -) 49 families / 61 participants / 12 fathers of children with CZS  -) participatory group intervention called “Juntos”  -) 10 sessions: ice-breaker activities, practical sessions, group discussions  -) observation checklist, participant observation  -) focus group discussions  -) interviews of fathers  -) evaluation of feasibility and acceptability of intervention | -) topics in groups: positioning/moving, eating, drinking, communication, play and early stimulation, everyday activities, community inclusion, disability rights  -) barriers to engagement of fathers: work commitments (provider of ﬁnancial support – self-awareness: practical and resource support) mothers as primary caregiver  -) stress, vulnerability: emotional/physical needs of child with disabilities, maintain family functioning; social isolation, stigma, mental health challenges, increased ﬁnancial/emotional strain; high risk of fathers abandoning their child  -) discussion and awareness-raising through media on Zika: less shame related to having child with CZS or similar conditions  -) after sessions: behaviour changes and conﬁdence in childcare, benefits: opportunity to share experiences; increased communication with child, learning practical skill (f.e. brushing teeth), improved knowledge | -) group format and content of “Juntos” well accepted  -) positive impact of father on child’s behaviour and development (school readiness, cognitive development, pro-social behaviours  -) better understanding fathers’ perspectives, needs and preferences within their context important to increase involvement in parenting interventions  -) changes in perceptions of childcare as “women’s work” |
| Duttine A., et al.  [30] | Brazil, 2020 | mixed-method study  6/2017 | -) need, relevance and use for post-Zika Brazilian context of similar structured family support programme to GTKCP (developed to educate and empower caregivers in childcare)  GTKCP = impact on the long-term health, wellbeing, participation of children with cerebral palsy; (practical, educational, peer-support and psycho-social aspects) | -) 30 families  -) 10-12 healthcare providers  -) case-control study  -) systematic review: needs of families of children with CZS and CP  -) Findings from social and economic Impact of Zika Study (usefulness of family support programme)  -) in-depth interviews  -) visit of institutions offering services to children with CZS | -) psychosocial aspects of caring for child with CZS or CP: anxiety, depression, stress, less life quality, lack of sleep of parents, ﬁnancial hardships; diﬃculties with transport and services, stigma  -) barriers: high healthcare needs of aﬀected children – frequent visits to health care services for conditions related to CZS (neurology appointments, physiotherapy etc), co-morbidities (chest infection, epileptic seizure); fragmented and uncoordinated, far distances, distrust, communication problems, diﬃculties for healthcare professionals addressing needs of these children and families; system for more severe cases, parents; response to CZS has more medical/therapy-based focus (rehabilitation available in specialised health centres in cities), no discussion of parent´s needs  -) information gaps: about child’s condition’ and appropriate institutions that the child can beneﬁt from  -) informal social networks between families to connect with each other, self-eﬃcacy for parents | -) GTCKP adapted to location and culture can be used as basis for family support intervention  -) more family support programmes in Brazil to ﬁll gap in Zika response additionally to clinical services and groups/ networks in post Zika context  -) adaption for diﬀerent age groups  -) more psychosocial, emotional support for caregivers, addressing social inclusion, respect to mental health |
| Kuper H., et al.  [31] | Brazil, 2018 | mixed-methods approach | -) ensure including people with disabilities in programmes  -) development of care support intervention targeting parents of children with CZS in Brazil  -) support families: providing psychosocial support, improving their skills in caring for their child optimally, connecting to available services | -) parent groups and their children with CZS  -) 10-11 sessions over 3 months, each session 3 hours  -) health promotion efforts with focus on improving knowledge,  skills and behaviours of parents | -) promotion of health and healthy lifestyles through behaviour change (nutrition, undertaking regular physical activity, engaging in vaccine and preventative health initiatives)  -) stress for families: poverty, financial strain through medical costs, lost income, risk of paternal abandonment and mental health issues for mothers  -) development of family support program: psychosocial support, improving skills to care for child, health seeking behaviour, sharing of experiences, addresses disability rights, guide child´s inclusion in school and health care, self-sustaining parent groups, emotional support activity, open and supportive discussion between parents in atmosphere of empathy and solidarity, empowers parents | -) poor health: poverty and inequality; people with disabilities more vulnerable  -) including people with disabilities in health promotion  -) parent/caregiver support programmes offer sustainable ways to improve skills and knowledge of parents and the health of children with disabilities, improve stigma/exclusion, poverty, carer distress |
| Ledogar R., et al.  [64] | Mexico, 2017 | descriptive | -) explain the Socialisation of Evidence for Participatory Action (SEPA) concept  -) examples of SEPA application in different countries and contexts  -) SEPA approach in the Camino Verde intervention in Mexico and Nicaragua  -) Centro de Investigación de Enfermedades Tropicales  (CIET) | -) random assignment of communities  -) house-to-house visits  -) facilitators  -) community volunteers (brigadistas)  -) use of evidence obtained from communities | -) SEPA: approach to production and use of evidence for health promotion and community development, includes social and cultural context, solution finding in dialogue with people  -) social components and cultural implications important  -) participatory action leads to behaviour change  -) tries to avoid pitfalls of social manipulation  -) cost-effective (increase efficiency for similar cost); community priorities  Effects of SEPA in this study:  -) “learning by doing”, “ownership effect”  -) repeated visits transform distrust to confidence  -) brigadistas (=”backbone”) motivate residents  -) facilitators (important role): community members, acceptable to community, initial contact, trust, gain access to households, recruite and work together with brigadistas on collective activities to eliminate mosquito breeding sites, provide training, raise and support responsibility of community for intervention  -) empowerment to interact with public authorities | -) SEPA approach appropriate for community mobilization in fighting vector borne diseases  -) applicable in various community and country conditions.  -) cost evaluation recommended, SEPA is cost-effective  -) need to develop and test community-based strategies  that include environmental management |
| Arostegui J., et al  [65] | Nicaragua, 2017 | description paper of a cluster randomised controlled trial  /feasibility study  -) feasibility  Study 2004-  2008, trial 2010  to 2013. | -) description of  Camino Verde intervention in Nicaragua  -) information on costs and discussion of location  -) collaboration with MoH’s National Diagnostic and Reference  Center (CNDR) | -) based on SEPA process  -) trial intervention  7/2011-12/2012  -) Facilitators  -) research team  -) biological, entomological, epidemiological and economic evidence  -) inclusion of saliva samples from children aged 3-9 before/after  dengue season in both  phases  -) baseline and follow-up surveys | -) dialogue with MoH and CIET: information exchange, joint action on mosquito breeding habitat (public places, natural environment, locations beyond community’s ability to control, recycling, tire repair shops, workshops, factories)  -) 3 activities to ensure participation in intervention:  (neighbourhood peer monitoring; neighbourhood responsibility, activities among neighbourhood brigades/ gatherings, common events)  -) household budget decisions with community leader, people became motivated to find solutions to decrease costs and think about handling mosquito problem  -) consent, trust, confidentiality: critical point in relationship between brigadistas and community members: initial interaction with residents  -) sharing evidence of mosquito populations in water containers with community leads to household and community actions in preventing spread of viruses  -) community empowerment: monitoring and evaluation activities by communities themselves | -) Camino Verde project registered reductions all vector control indices  -) elimination of breeding habitat part of everyday household cleaning (aim)  -) demonstration of positive impact of pesticide-free community mobilisation on virus infection in children  -) evidence based on living conditions, leads to more mobilisation than use of didactic material  -) application of similar process under specific circumstances in other parts of the world |
| Morales-Perez A., et al. [66] | Mexico (Guerrero), 2017 | descriptive study | -) engagement of intervention teams with  communities to share information and to plan prevention activities in the Mexican arm of Camino Verde trial  -) activities at household and community levels, including interactions with service providers | -) 90 coastal communities  -) SEPA process  -) 4-point protocol for communities to identify preventive actions after discussion (stakeholder discussion, co-design of prevention activities,  training of community  volunteers (peer evaluation)  -) 182 discussion groups  -) 45 intervention clusters  -) 4 week training of facilitators  -) house-to-house visits  -) clean-up campaigns  -) intervention and control communities  -) government vector control programme continued (temephos) | -) perception: mosquito control considered to be government responsibility  -) barriers: conflicts between neighbours, lack of water supply/ services, insecurity, land invasions by squatters, other health problems, dengue not prior to most; challenging security conditions: social violence eliminated local community structures, no attendance to meetings because concern about security  -) household level: saliva sample tests, brigadistas shared evidence/ inspection of breeding habitat/tools (laminated cards)  -) community level: biological control using fish, mid-course peer evaluation (exchange between brigadistas from different communities), working with schools (became centres of efforts to vector control, inducing activities outside school)  -) success: facilitator training 4 weeks before intervention (community authorship); diversity of community involvement & dialogues, house-to-house visits for mobilisation; rise of new leadership in some communities (more confidence); triggering volunteerism, increasing social capital (household awareness led to joint clean-up campaigns in public) | -) brigadistas/ facilitators as link between community members and public services  -) collaboration with local schools recommended (recycling programme, collective benefit)  -) impact of community empowerment and ownership of initiative  -) building authorship and ownership required (not copying preventive activities) for success in other places |
| Weldon T. C., et al. [53] | Peru, 2018 | qualitative study,  6/2017 | -) knowledge, attitudes and prevention behaviour related to Zika virus | -) Iquitos (amazon basin)  -) 46 women age 20-35  -) invitation by local field worker near homes  -) 6 focus group  -) use of health belief model to inform focus group discussion guide  -) travel costs taken by research team | a) knowledge: symptoms (rash, mild fever, headache, muscle pain, red eyes) and mosquito transmission; few knowledge about neurological disorders, sexual transmission and prevention options; existing public health campaigns to prevent transmission via radio, pamphlet, billboards; health education/ information: not always provided well, women were receptive  b) Perception: rare healthcare seeking, awareness of Zika risks, but remaining at home no option, severity: lowest ranking of Zika (mild symptoms, no microcephaly cases), concern for pregnant women and foetus, women´s responsibility: condom use as challenge (partner disputes), unintended pregnancies  c) prevention practices: general household cleanliness, mosquito nets, repellents, reducing breeding sites, treatment: refreshments (juices from local fruits, malba leaves) paracetamol  -) no laboratory testing, symptoms lead to diagnosis | -) knowledge gaps in prevention practices, more education/ information needed  -) peer education: people are participants in the process (not objects or recipients), identify problems and solutions  -) dengue research for 25 years, most health campaigns related to Dengue virus (emergency fumigation, larvicide application, eliminating water containers)  -) report of Zika in 11 of 25 Peruvian departments |
| Iguiñiz-Romero R., et al.  [35] | Peru, 2020 | rapid qualitative assessment  design,  2 Phases:  8-9/2017  1-2/2018 | -) focus on early  healthcare response to Zika in Peru  -) perceptions of frontline personnel (midwives, nurses), their  role in prevention of Zika virus and management of CZS | -) Piura, Peru  -) personal invitations  -) focus groups  -) in-depth interviews,  -) focus group questionnaire: interventions in  context of Zika virus | a) knowledge: High awareness but low knowledge and preventive practices; lack of training (midwives received training on Zika-related care during early response, nurses not), rare knowledge about CZS  b) perception: benign nature of Zika compared with Dengue, no cases of microcephaly; low perception of Zika virus risk might reflect attitudes; belief Zika has periodic activity like dengue  c) barrier: limitation of primary health system (access for prenatal visits) targeted women of reproductive age  -) health personnel at primary health care centres share cultural aspects with communities | -) more cooperation between nurses and midwives, engagement as primary health care providers  -) overlapping epidemics (Zika + dengue): difficult to differentiate  -) more long-term surveillance, diagnostic resources, management of ZV and CZS, training |
| Sousa CA, et al. [54] | Brazil, 2018 | qualitative, descriptive-exploratory study,  3-5/2017 | -) knowledge, perceptions and care practices of women with Zika virus infection during pregnancy  -) subproject of a matrix project by Universidade Estadual de Mato  Grosso | -) basic health units, Mato Grosso state  -) 10 women age 18-36 (Zika diagnosis during  pregnancy)  -) open interviews | a) knowledge: little information from health services rare knowledge; Internet, TV, mass media information campaigns  b) perception: more fear and concern about baby than own well-being, fear of transmitting infection to baby during pregnancy, belief that virus always causes malformations, non-biological consequences to infected women (depression); medical care seeking because of body changes, no expectation of Zika  c) prevention: education level influences diseases prevention, less preventive measures during gestation after some time, lack of scientific evidence that Zika is associated with microcephaly  -) controversy between recommendations for high-risk-prenatal care and follow-up of pregnant women by attending professionals | -) lack of publications in Portuguese language difficult for healthcare personnel update  -) deficit of medical and public health services guidance (rare data collection after 2 years of first Zika case)  -) better prenatal care  -) future research in different contexts, exploring experiences and care practices of pregnant women infected with ZV |
| Martaleto JL, et al. [36] | Brazil, 2017 | qualitative study,  2015-2016 | -) responses of vulnerable women in pregnancy intentions and in sexual and contraceptive behaviour  -) women´s desires, behaviours, healthcare access and use during the first 18 months of the Zika epidemic  -) overview of the Brazilian context | -) two regions of Brazil (Belo Horizonte and Recife)  -) 114 women, age 18-49  -) 8 focus groups, 6-8  women  -) low and high socioeconomic status  -) neighbourhood recruitment  -) interview protocol,  open ended questions,  same interviewer  -) transport costs reimbursement (15$) | a) Brazilian context: i) many unintended pregnancies, especially if low socioeconomic status (if low more children, earlier) ii) ZIKV epidemic first in Northeast Brazil: lower economic development, high temperatures, stagnant water, sanitation problems, higher fertility rates; good economic development in Southern areas, iii) some seek abortion (illegal), iv) awareness of consequences of ZIKV  b) risk perception: i) fear of contracting ZIKV, postpone pregnancy 2-3 years (low- and high-SES women); age is key factor for postponement ii) more reproductive control in high SES women (low SES more vulnerable) iii) belief that more abortions as consequence of epidemic iv) prevention: repellent, long-sleeve shirts, winter months (low mosquito prevalence); Reproductive behaviours: i) majority of low-SES group reinforce/ start with contraception due to epidemic; high SES women: continue contraception, very few unintended pregnancies, ii) abortion: illegal; medical pills, teas; medical/surgical  c) barriers: i) availability at public clinics (pill, injection, condom), stigma, privacy problem, gossip (same community as clinic staff). private clinics for high SES women (larger variety: IUD, vaginal ring); sterilisation; ii) bargaining with partner, men do not worry about Zika. iii) time consuming, price (valid prescription, picture ID needed to pay less) iv) safe abortions not accessible for low SES women | -) socioeconomic status and ZIKV prevalence influence women´s pregnancy decision  -) inequalities in reproductive healthcare access  -) improving access to reproductive health care services decreases longer-term consequences of health inequalities  -) reduce barriers to contraception (availability of long-acting reversible contraception, contraceptives in all pharmacies)  -) more education and guidance in sexual healthcare |
| Borges ALV., et al. [8] | Brazil, 2018 | qualitative study,  8-9/2016 | -) knowledge, pregnancy attitudes, contraceptive practices in relation to the Zika virus outbreak in Brazil  -) evaluation of attending reproductive  healthcare services  -) evaluation of  women´s pregnancy  and contraceptive decision in context to Zika in Brazil | -) Aracaju, north-eastern Brazil  -) 526 women, age 18-49  -) 19 primary health care services  -) approach in the waiting area  -) interviews  -) interviews by nurses or psychologists | a) knowledge: high awareness about CZS, low about sexual transmission, independent from social background, age  b) prevention: minority of pregnant women use condom to prevent ZV, contraceptive use depends on healthcare provider´s advice on avoiding pregnancy, majority not advised to delay pregnancy or prevent perinatal transmission; low use of LARC (almost non-existent, focus on short acting methods)  c) perception: few women changed pregnancy decisions in response to Zika outbreak, not related to social status; influence on abortion decision-making  d) barrier: poor access to information about prevention of ZV transmission; emergency contraception in private pharmacies; inadequate family planning response to Zika, health system responses related to pregnancy occurrence; 60% pregnancies unintended in study population | -) training of primary health professional teams (focus on diagnosis and clinical management)  -) family planning services and prevention of perinatal transmission; evaluating pregnancy intentions should be included in routine care |
| Anderson JE., et al. [22] | Brazil, 2020 | qualitative study,  6/2016 | -) experiences and barriers to Zika-related  healthcare access  -) knowledge in high-risk low-income urban population  -) partnership of scientists with community-based NGO | -) Fortaleza  -) 37 women > 18 years  -) snowball recruitment  -) invitations by NGO  -) interviews, open ended  and closed-ended questions on social factors | a) knowledge: low awareness of Zika epidemic maybe due to poor access to primary healthcare, 22% aware of Zika consequences, 50% know Zika transmission by mosquitos  b) prevention: personal responsibility: most common prevention strategy is cleaning - health campaign information on tv, radio, flyers; few use condoms regularly, use of contraception not reported as Zika prevention strategy  c) barrier: poor sanitation and healthcare access/ unable to identify medical or social services, poverty, lower education; Response to Zika in Brazil at state level restricted: i) insufficient funding ii) political instability iii) limited administrative capacity  -) business as usual concerning mosquito-borne disease  -) low-income women more unwanted pregnancies  -) long-term consequences of CZS worse due to inequitable healthcare access | -) Zika prevention campaigns 2015 und 2016: focus on mosquito reduction education, not on sexual transmission  -) more family planning services (condom, birth control)  -) Zika cases decreased but still long-term risk; more infants with CZS  -) more communication of differences between Zika and Aedes-borne viruses  -) future research: appropriate exploration culturally and linguistically |
| Diniz D., et al. [23] | Brazil, 2020 | qualitative study,  11/2017-7/2018 | -) examine barriers for young women living in Zika affected areas and seek sexual and  reproductive health  (SRH) care services  -) attitudes towards  SRH, needs and available services | -) 3 Zika affected towns in north-eastern Brazil  -) 3 health care facilities  -) 22 women aged 14–24  -) Sentinel Sites Evaluation (SSE) activities (by WHO Human Reproduction Program)  -) invitation by health  workers, women seeking sexual and reproductive  health services  -) interviews | a) knowledge: Zika virus not considered as health concern, low awareness of sexual transmission, lack of information: i) knowledge gaps about contraceptives use (how to use) ii) misconceptions on contraceptive methods adverse effects and effectiveness; microcephaly recognized consequence of ZV; little information and barriers to access SRH might be reason for misconception; low education level  b) perception: all participants experienced unwanted pregnancy, young age of first pregnancy, reason for leaving school  c) barriers: Communication failures, unawareness about Zika and consequences (inability to provide adequate information), long waiting time for appointments, lack of desired birth control methods (LARC), access to family planning, | -) social and gender inequalities increase vulnerability of women and families  -) better access to health information and family planning services (age, biopsychosocial context to provide SRH and reduce unwanted pregnancies)  -) more focus on women in public health response |
| Mendoza C., et al. [24] | Colombia, 2020 | qualitative study,  2018 | -) knowledge and concerns of pregnant  women in context of Zika and transmission  -) perceived importance of Zika  -) personal protective technologies (PPT)  -) routine and desired mosquito control activities | -) Cali and Villavicencio  -) 143 (pregnant) women, 14 men, age 18-45  -) recruitment through  organized groups (pregnant women groups,  groups of SRH programs, childcare groups), telephone  -) 19 focus group discussions with female participants,  -)14 semi-structured interviews with men | a) knowledge: good knowledge of mosquito transmission, rare about sexual transmission, shortfalls of consequences associated with Zika (malformations)  b) perception: trust in community health workers, safety fears/concern about health risks of repellent chemicals during pregnancy, concern about costs of repellents in lower SES groups; disparity between diseases Dengue and Zika (concern higher for Dengue) more familiar with Dengue because of former epidemics (Haemorrhagic Dengue 1990)  c) prevention: repellent clothing; larval source reduction only preventative action for Zika, health centres best source of information on bite-reduction; easy use of PPT enables | -) engagement of key users in communities  -) educational campaigns during antenatal service (Zika risks during pregnancy, repellent safety)  -) collaboration with stakeholders, involving societal, economic and political context |
| Gomez M.H., et al. [26] | Colombia, 2020 | qualitative study  9/15-12/16,  4/18-6/18 | -) explore perceptions and experiences of  pregnant women in accessing healthcare services during the epidemic in Colombia during 2015-2016 | -) Villavicencio  -) women diagnosed with Zika  -) recruitment through National public health  surveillance System  SIVIGILA, municipality´s records  -) 6 participants  -) semi-structured interviews  -) topics: maternity care, family planning, usage history, maternity expectations, cultural beliefs | a) knowledge: basic information about symptoms (fever, malaise, cutaneous rush; consequences for foetus: microcephaly; transmission: mosquito, attending gynaecologists and nurses but no comprehensive provision of information about Zika  b) prevention: repellent use, bed nets, postponing pregnancy  c) barriers: delayed medical appointments (availability, limited supply in municipality; prenatal check-ups; delayed access to prenatal laboratory testing and ultrasounds, delayed test results, difficult access to healthcare services for new-borns with suspected microcephaly (limited specialized physicians), out-of-pocket payments (medical consults, laboratory tests); administrative procedures, authorization denials; lack of continuity of care, gaps in knowledge of existing guidelines during pregnancy | -) improvement of health education by healthcare professionals  -) focus of SRH on women age 15-49  -) more communication between stakeholders |
| Casapulla SL, et al. [37] | Ecuador, 2018 | cross-sectional design (pilot  study);  5/2016 | assessment of Zika virus-related knowledge  and attitudes among adults in Ecuador  . | -) small cities and rural areas in the Amazon and Andes region  -) 181 participants, av.  age 33  -) two visits in communities before beginning of the study  -) group settings  -) information about  ZIKV and HIV (1 page-pictogram, “me quedo  frio”)  -) condom distribution,  donation to public health clinic  -) health-belief- model | “Zika Triad”: risk reduction, dealing with information gaps; susceptibility to Zika, consequences, way of thinking  a) knowledge: 53% from tv; majority knew about mosquito transmission, low (8%) about sexual and in-utero transmission; knowledge based educational level  b) perception: low perceived susceptibility (HBM) - low rating of own risk and circumstances for getting disease; misconceptions: day/night biter; belief that mosquito bite prevention is enough, belief in vaccine protection against ZIKV though no existing vaccine, diseases perceived as severe when visible symptoms and consequences  c) barriers: condoms available but not used (catholic, cost)  -) authors known and respected in community due to former visits | -) need for educational initiatives about transmission and prevention of ZIKV  -) increase access to contraception, bug spray and protective clothes  -) exploration of reasons behind perceptions of severity |
| Fritzell C. et al.  [60] | French Guiana, 2017 | cross-sectional phone survey,  6/2016 | perceptions, knowledge and behaviours in Zika context | -) 1129 participants  -) phone recruitment  -) interviews during the day, standardized questionnaire | a) knowledge: symptoms fever, myalgia, headache, arthralgia; 80% mother-to-child, 55% sexually transmission awareness of Zika, communication media available to all subgroups (tv, radio, posters, leaflets)  b) perception: more serious health threat than other mosquito diseases, easier to avoid than Dengue; behaviour variety among social groups, geographic areas and gender (lower educated women: more worries, less protective behaviour), women addressable to awareness campaigns  c) prevention: eliminating water storage, bed nets, covering storage containers; insecticide spraying, use of vaporizer for outdoor insecticides | -) health institutions should reach less advantaged women, visit mother and child protection centres  -) more public information, emphasize importance of protective actions during outbreak  -) surveys |
| Shaw R., et al. [27] | Dominican Republic, 2019 | multi-site, mixed-methods, cross-sectional study,  3/2016 and  3/2017 | -) assess knowledge of Zika virus, use of contraceptives and information sources in rural communities in the Dominican Republic  -) report findings of 2017 pilot study  -) collaboration Des  Moines University and Timmy Global Health | -) 4 rural communities  -) 90 participants  -) 12-question survey  -) community outreach  -) local hcp instructed  residents about free health care by research team before outreach  -) provision of health  care services over 4 days  -) evaluation interview  for team members | a) knowledge: deficits about virus; but most participants think ZIKV present, some believed ZIKV curable (problem for public health prevention efforts)  -) low knowledge about sexual transmission  b) prevention: rare use of LARCs, limited protection against mosquitoes (no window screens, no long-sleeved clothing)  c) barriers: socioeconomic factors block prevention of infection  -) conducted in impoverished and underserved areas, not representative of whole nation | -) engagement of stakeholders  -) further community engagement campaigns  -) recommendation for use of contraceptives and education about vector-borne diseases  -) more surveys to assess public health education |
| McDonald A.J., et al. [56] | US/Mexico, 2018 | qualitative study,  10-11/2016 | -) awareness and  knowledge of ZIKV  among pregnant; prenatal care for women  -) information sources | -) border counties US-Mexico  -) healthy start program  -) pregnant women or of childbearing age  -) 326 participants  -) recruitment: caseworkers, during home  visits or clinic, healthy  start appointments  -) interviews with open-ended questions | a) knowledge: two ways to prevent infection, familiar with ZIKV on the US-Mexico border, information sources: health care providers (women more knowledgeable about prevention and testing). TV least helpful, radio  b) prevention: gaps in use of condoms or abstaining from sex  c) barriers: access to health care providers (<50% women mentioned hcp though being in healthy start program)  -) local and travel-associated ZIKV cases  -) 5 healthy start programs in the region; PRAMS pregnancy risk assessment and monitoring system | -) CDC´s PRAMS survey added 12 standard questions about ZIKV  -) More coordinated health education, emphasizing pregnancy postponement and sexual transmission of ZIKV  -) more focus on education during meetings in clinics or home visits |
| Tirado V., et al. [38] | Colombia, 2020 | qualitative study,  2017 | -) experiences of ZIKV infection during pregnancy by affected  women and the influence on their family relationships and future family planning  -) collaboration National Public Health Surveillance System  SIVIGILA and Secretariat of Health, Medellin | -) Medellin  -) SIVIGILA records  2015-2018  -) 19 women, age 18-45, confirmed Zika infection during pregnancy  -) semi-structured interviews | a) knowledge: tv, radio, healthcare providers, doctors, workshops in community/school, preference of educational sessions (different mosquitoes types, symptoms); aware of mosquito borne diseases, impact of infectious diseases more comprehensible if infection happened in community, no knowledge of sexual transmission  b) perception: concerns about consequences after diagnosis, potential social stigma, after ZV diagnostic change in relationship between father and child; abortion: legal in Colombia in case of rape, incest or to protect woman´s health, influenced by healthcare providers, religion, social attitude  c) prevention: removal of breeding habitat, personal problem with wearing long sleeves all day  -) diagnosis: mainly during 3^rd^ trimester, all 19 births suspected having CZS; unclear feelings, co-infection/cross-reactivity with dengue | -) challenges: medical care; economic burden, uncertainty about CZS, limited professional knowledge, social isolation and stigma  -) need for better healthcare service after birth, support/guidance after diagnosis  -) establishing family planning services and long-term programs for children with CZS  -) consider male involvement in sexual and reproductive healthcare |
| Bailey DB.Jr., et al. [74] | Brazil, 2018 | essay | attention to lifelong consequences of CZS for families, long-term needs of families and social burden | -) find risk and protective factors concerning children´s development and family well-being  -) understand and maximize family adaption  -) challenges for families of children with CZS | -) family-oriented approach to support: providing information about CZS, surveillance to identify needs, facilitate access to support systems, interventions to support child health, child development and positive family adaption  -) implications of CZS, 4 features: i) medical complexity and severity, lifetime caregiving and economic burden, ii)) uncertainty about consequences of CZS iii) limited professional knowledge about course of disease or treatment options iv) social isolation, few social or community supports, stigma | -) engagement and support of families in future as part of health care |
| Gurman T., et al.  [75] | Dominican Republic, 2020 | qualitative study,  4/2018 | -) influence of gender´s role on Zika prevention behaviours in the Dominican Republic  -) cooperation research team with USAID and local NGO partnership | -) peri-urban municipality  -) pregnant women,  women of reproductive age, male partners age  18-30  -) 88 participants  - 12 focus groups  - 8 in-depth interviews  with men  -) discussions (by facilitators)  -) participant´s feedback on outcomes | -) prevention: condom use unnecessary during pregnancy; infidelity insinuations of condom use during pregnancy (fear of being accused by partner); some men active in household during pregnancy  -) prenatal care: opportunity to engage men and women in discussions about men’s role during the visits (neutral surroundings)  -) Zika prevention: expand men’s willingness helping partner, see co-responsibility in preventing infection  -) involvement of men in vector control behaviour adapted to local context (condom use to protect family)  -) manage sensitive conversations among couples with respect to cultural context, intimate partner violence prevalent, increasing machismo in DR | -) more welcoming atmosphere at healthcare services, education about prevention  -) dealing with role of gender in Zika prevention strengthens programs  -) integrating of gender issues in local context, not exploiting existing gender roles (protector) |
| Darney BG, et al. [28] | Mexico, US (Texas), 2017 | commentary,  2017 | -) role of health system factors in access to contraception regarding  Zika, Texas and Mexico  -) contraceptive access efforts in Puerto Rico | -) commentary  -) analysing two case studies  -) illustration of health inequities and population health  -) comparing efforts in Puerto Rico to improve contraception access | a) prevention: “stopping” vs “spacing” = after reaching desired family size use of permanent contraception (sterilization); adolescent births are government priority issue (abortion: 1^st^ trimester legal since 2007, free of charge in public sector); focus of response to Zika on vector control, no advice for avoidance of pregnancy, no family planning initiatives to support  b) barriers: obstacles in pregnancy decision especially for nulliparous, adolescent, indigenous women = group with lowest knowledge of options and most limited access to services (emergency contraception, abortion). highest risk for Zika but least able to delay pregnancy | -) more efforts to remove barriers of contraception access to prevent unwanted pregnancy in Zika context  -) links between mosquito-borne disease, poverty, access to health services in general and reproductive healthcare  -) Zika virus shows weaknesses in/of health systems |
| Bardach AE, et al. [67] | Argentina, 2019 | systematic review | collect information on effectiveness, cost-effectiveness of vector control strategies (Ae. aegypti in Latin America and the Caribbean (LAC)), realization and reported experiences | -) database: Medline, Embase, Central, Socindex, Lilacs, websites of WHO, NGOs, Google Scholar  -) systematic search  -) PRISMA guidelines  -) 75 included records from 2000-2016  -) meta-analysis of 9 cluster randomised clinical trials  -) part of wider mixed qualitative and quantitative research | -) data from Brazil, Argentina, Cuba, Mexico, Peru.  -) IVM in combination with community engagement effective, long-term outcomes with health education, reduction in larval indices, reservoirs and mosquitoes  -) education and communication (IEC) strategy, surveillance programs, school programs or training of community leaders, community empowerment initiative targeting participatory processes  -) prevention: reduction of entomological indices: health education, community engagement, use of insecticide-treated materials, indoor residual spraying (low certainty of evidence), container management; lethal ovitrap mass interventions; no relevant reduction but high acceptance for insecticide materials: bed nets/curtains, indoor residual spraying, insecticides in breeding sites, surveillance to improve indices | -) community engagement and education: community mobilisation programs effective to reduce indices  -) lack of evidence on effectiveness of vector control but some interventions showed moderate success  -) longer duration of interventions and sustainability |
| Alvarado-Castro V., et al.  [68] | Mexico, 2017 | systematic review,  2013, 2015, 2016 | -) identify cluster randomised controlled trials of interventions to control Aedes aegypti  -) evidence of effectiveness of different control measures in reducing Aedes aegypti | -) database Medline, Ovid, BVS, Lilacs, Artemisa, Imbiomed, Medigraphic  -) systematic research  -) 3 searches  -) CRCTs published  1/2003-10/2016  -) 18 articles (9 community mobilisation, 8 chemical control, 1 biological control)  -) impact on household index (HI), container index (CI),  Breteau index (BI) (= standard entomological indices) | -) chemical control interventions: insecticide-treated window, door screens or curtains, combined with insecticide-treated water container covers, temephos or spinosad treatment; education/clean-up and visits as “control” condition  -) biological control interventions: avoiding chemicals but no large scale application  -) community mobilisation and participation interventions: most effective; household and community level, dialogue with local stakeholders to discuss and plan activities, engaging community members in preventive activities; home visits to help with breeding sites removal; partnerships with local institutions to improve services (garbage collection), involving schools and women; implies changes in attitudes and behaviour, motivation, sustainability: evaluation of evidence and co-designing interventions with respect to local conditions and culture | -) community mobilisation programmes most effective intervention to reduce entomological indices  -) chemical vector control prevention efforts by government combined with community mobilization recommended |
| Rodrigues SA. [61] | Brazil, 2019 | cross- sectional  study,  3-5/2015 | -) investigation of psycho-cultural perspectives regarding family quality of life among Brazilian families with children who have intellectual disability  -) impact of severe or profound intellectual disabilities on the FQoL dimensions | -) Sao Carlos  -) 15 women of children with severe intellectual disabilities, age 15-21  -) recruitment through Public Medical Genetics Service, Association of Parents, Friends of exceptional children of Sao Carlos  -) semi-structured interviews  -) Family Quality of Life Survey 2006 framework | -) public health system (SUS) “Healthcare Network for the Disabled”: health inequity; restricted access to healthcare services and support; transportation and recreation; difficulties in primary health care infrastructure, delays in medical appointments, no beds  -) 2014 National Policy on Comprehensive Care for People with Rare Diseases established: programme for people with intellectual disabilities in the SUS  -) mythification of mothers=main person in childcare, interrupt career, results in financial difficulties, isolation, low self-esteem  -) intense relationship between mother and child with intellectual disability, less attention for “sane” child  -) stress: difficulties in obtaining information and access to health services, helplessness due to perception of low quality of care; social stigma: social interactions in community difficult; religious coping | -) improvement of emotional, psychological care and access to health care (lack of professional support especially in public facilities)  -) considering cultural context (social stigma, gender issues)  -) establish family support groups, empowerment through exchange with other affected families |
